# Incidence and Risk of Alzheimer’s Disease in Individuals with Type 2 Diabetes: A Systematic Review and Meta-Analysis

**DOI:** 10.64898/2026.07.08.26357542

**Authors:** Thi Thu Huong Nguyen, Asa Auta, Emmanuel Agada David, Chinedu I. Ossai, Victory Olutuase, Moulinath Banerjee, Yun Zhao, Davies Adeloye, Gavin Pereira, Emmanuel O Adewuyi

## Abstract

**Background:** Epidemiological evidence links type 2 diabetes (T2D) to an increased risk of dementia, including Alzheimer’s disease (AD). However, previous syntheses often combined heterogeneous diabetes and dementia definitions and have not comprehensively quantified AD incidence among individuals with T2D. We aimed to estimate both the incidence of AD among individuals with T2D and the association between T2D and AD using studies with well-defined T2D and AD outcomes.

**Methods:** We systematically searched MEDLINE, CINAHL (via EBSCO), Embase (via Ovid), and Scopus from inception to April 2026 for studies investigating the incidence of AD among individuals with T2D or the association between T2D and AD. Data were pooled using random-effects models and presented as incidence rates and adjusted relative risks (RRs) with 95% confidence intervals (CIs).

**Results:** Of the 9,430 articles identified, 40 studies involving 27,102,559 participants were included. Twenty-three studies contributed incidence data, and 26 reported adjusted relative risks (aRR). The pooled incidence of AD among individuals with T2D was 4.71 per 1,000 person-years (95% CI 3.31–6.71). T2D was associated with an increased risk of AD (aRR 1.53, 95% CI 1.38–1.70). Subgroup findings were generally consistent, results were robust in sensitivity analyses, and no publication bias was detected.

**Conclusions:** This study provides a comprehensive quantification of the AD burden associated with T2D by focusing on well-defined AD and T2D outcomes and advancing the field beyond prior broad dementia syntheses. Integrating incidence and relative risk estimates clarifies both the absolute and relative burden of AD in T2D and extends previous syntheses that primarily emphasised relative risk. Individuals with T2D experienced approximately five AD cases per 1,000 person-years and a 53% higher risk of AD, supporting the rationale for integrating cognitive risk prevention into diabetes care.

## Background

Alzheimer’s disease (AD), the most common cause of dementia, represents a growing global public health challenge. Worldwide, dementia affects more than 57 million people, with Alzheimer’s disease accounting for approximately 60–70% of cases, and the number of affected individuals is projected to increase substantially over the coming decades [1, 2]. Recent global estimates indicate marked increases in dementia prevalence, mortality, disability, and healthcare costs, driven largely by population ageing [2, 3]. The burden is evident across regions [4, 5]; for example, dementia has recently surpassed ischaemic heart disease as the leading cause of death in Australia [6]. These trends highlight an urgent need to identify modifiable risk factors that may contribute to dementia, particularly AD.

Type 2 diabetes mellitus (T2D) is one such candidate. T2D has reached epidemic proportions globally, affecting more than 590 million adults and accounting for over 90% of diabetes cases worldwide [7, 8]. Given that T2D and AD are both age-related disorders with overlapping cardiometabolic risk profiles [9], there has been a sustained interest in whether T2D independently confers risk for AD. Observational studies have consistently linked diabetes with both cognitive impairment and dementia, and multiple meta-analyses reported an increased risk of dementia among individuals with diabetes [9–14]. However, findings specific to AD are more variable and generally more heterogeneous than those reported for dementia overall [11, 15, 16].

Biological evidence supports the plausibility of an association between T2D and AD. Insulin and insulin-like growth factor-1 signalling influence neuronal survival, synaptic plasticity, and amyloid-β metabolism, and impaired central insulin signalling has been described in AD [17–19]. In parallel, the metabolic disturbances characteristic of T2D, including hyperglycaemia, oxidative stress, dyslipidaemia, and inflammation, may contribute to cerebrovascular injury and neurodegeneration [19–21]. Yet clinicopathological studies suggest that diabetes is more consistently associated with cerebrovascular or mixed brain pathology than with classical AD pathology alone [22–24], highlighting the need to distinguish AD from other dementia subtypes in epidemiological research.

Despite a longstanding interest in the association between diabetes and AD, important gaps remain in the evidence synthesis literature. Many previous reviews often pooled heterogeneous diabetes definitions or combined distinct diabetic exposures [11, 13–15, 25, 26], despite well-established aetiological differences between diabetes subtypes [27, 28]. Similarly, outcomes have frequently been reported for all-cause or incident dementia, whereas AD-specific outcomes were not always primary or consistently reported [13, 14, 25]. Consequently, existing syntheses provide limited insight into the specific relationship between T2D and AD. In addition, prior reviews have largely focused on relative risk estimates and have seldom quantified the incidence of AD among individuals with T2D. Although the evidence base has expanded substantially over the past decade, including large population-based cohort studies across diverse settings, no systematic review or meta-analysis has synthesised this growing literature while jointly evaluating both the absolute burden of AD (incidence) and its relative risk associated with T2D.

To address these gaps, we conducted a systematic review and meta-analysis of observational studies evaluating T2D and AD. Our objectives were to estimate the incidence of AD among individuals with T2D and to quantify the association between T2D and AD using adjusted relative risk estimates. We restricted inclusion to studies with well-defined T2D and AD and applied uniform inclusion and extraction criteria. This approach enabled a comprehensive and biologically informed assessment of the absolute and relative burden of AD associated with T2D, with clear clinical interpretability.

## Methods

### Information sources and search strategy

The study protocol was registered in the PROSPERO - International Prospective Register of Systematic Reviews (CRD420251231648).

Following the PRISMA 2020 guideline [29], we systematically searched the MEDLINE, Embase® (via Ovid), CINAHL (via EBSCO), and Scopus databases for articles published from inception to April 2026. The search strategy employed Medical Subject Headings (MeSH), truncations, and keywords along with their synonyms, including Alzheimer’s, Type 2 Diabetes Mellitus and observational studies, which were combined using Boolean operators (Table 1S – Supplementary materials). Additional articles were identified by Google Scholar citation tracking and checking the reference lists of eligible studies.

**Table 1:**
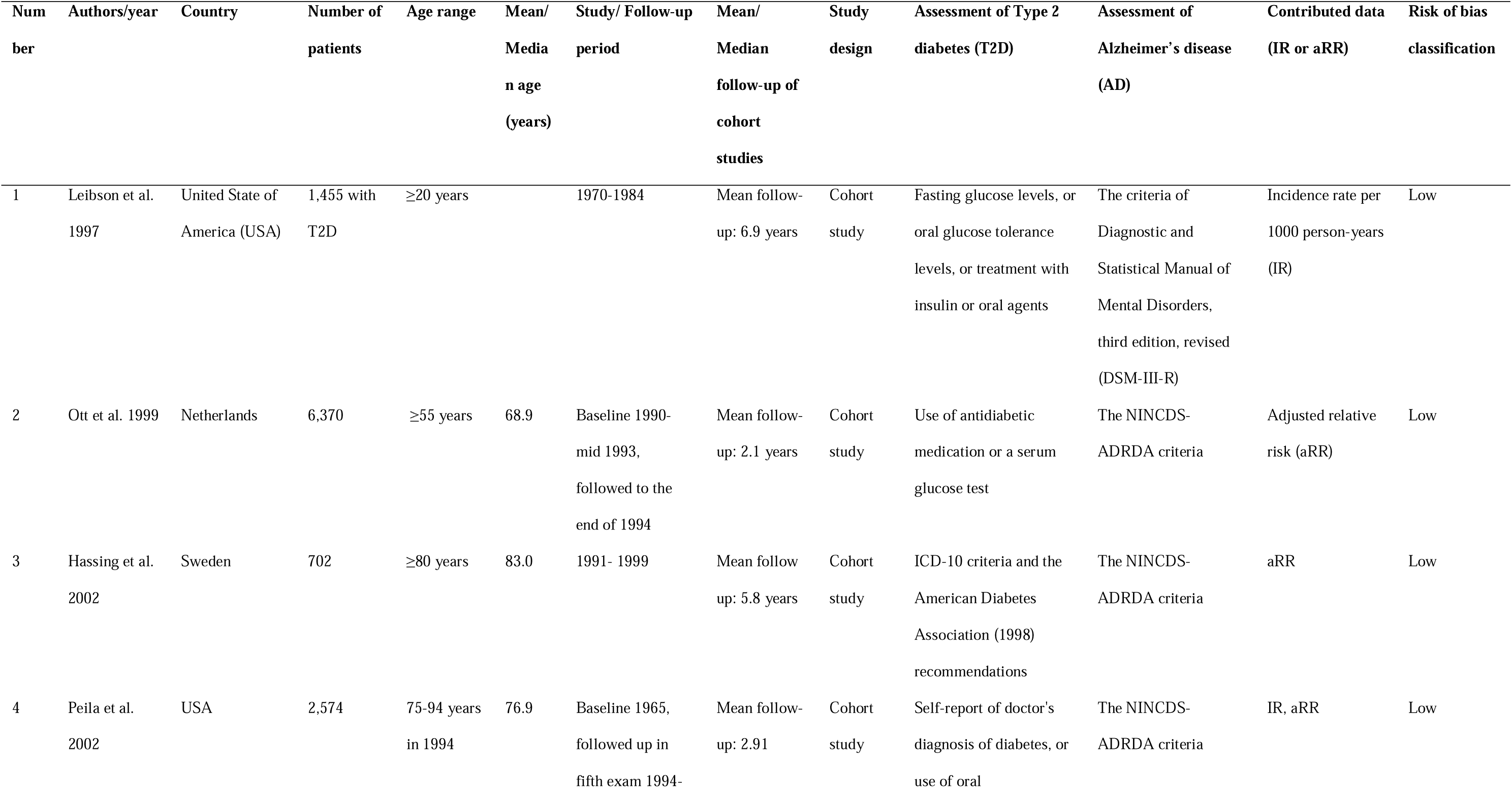
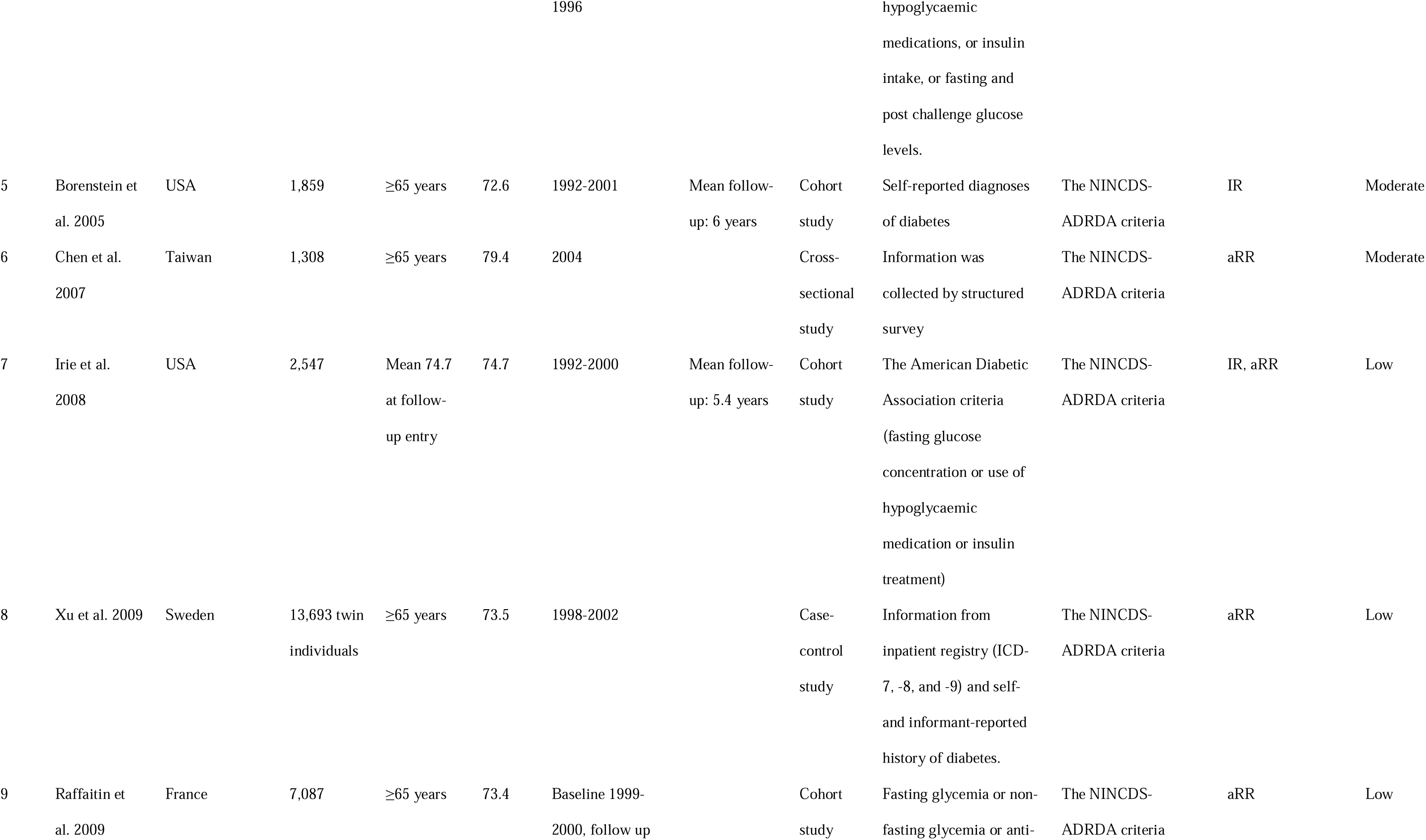
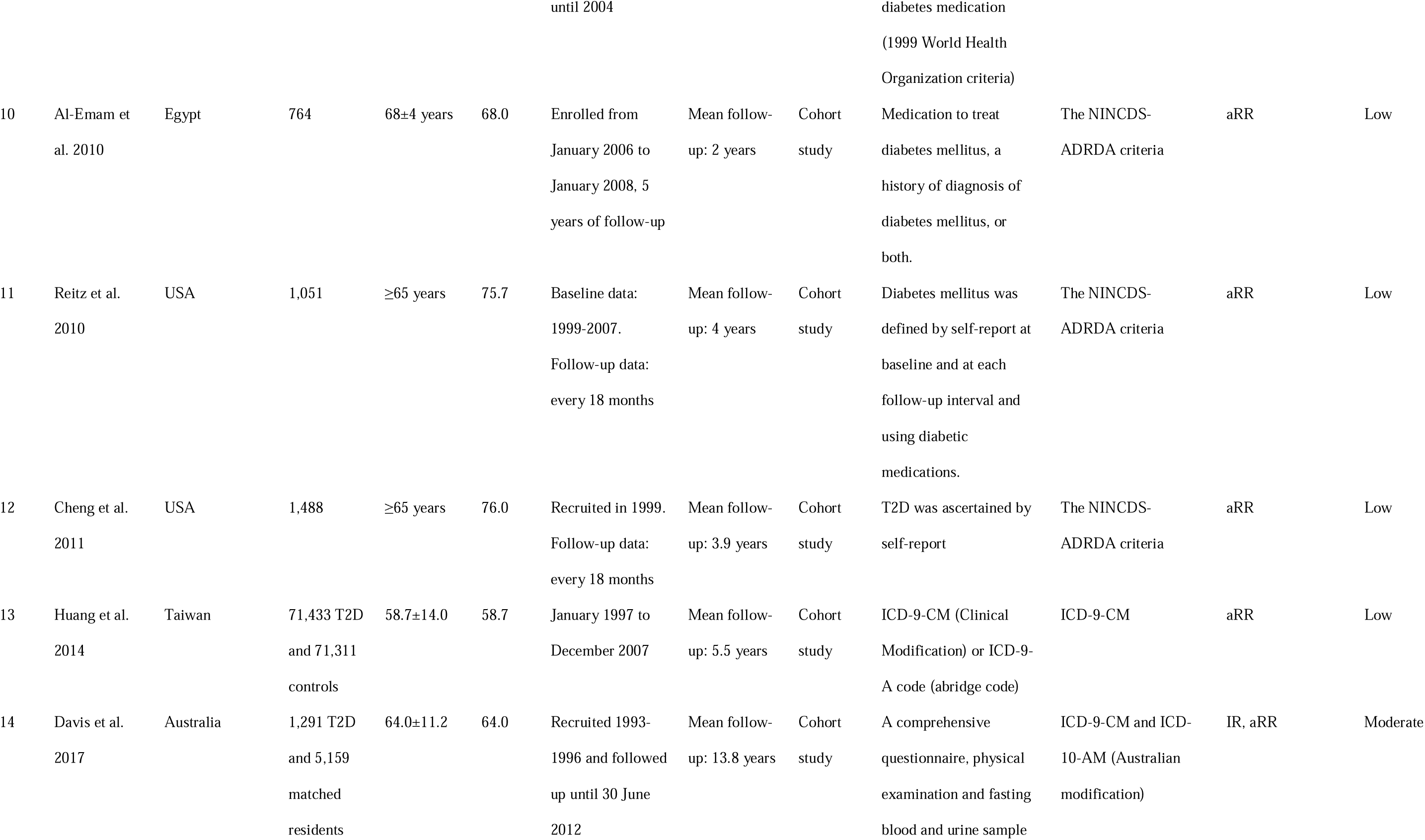
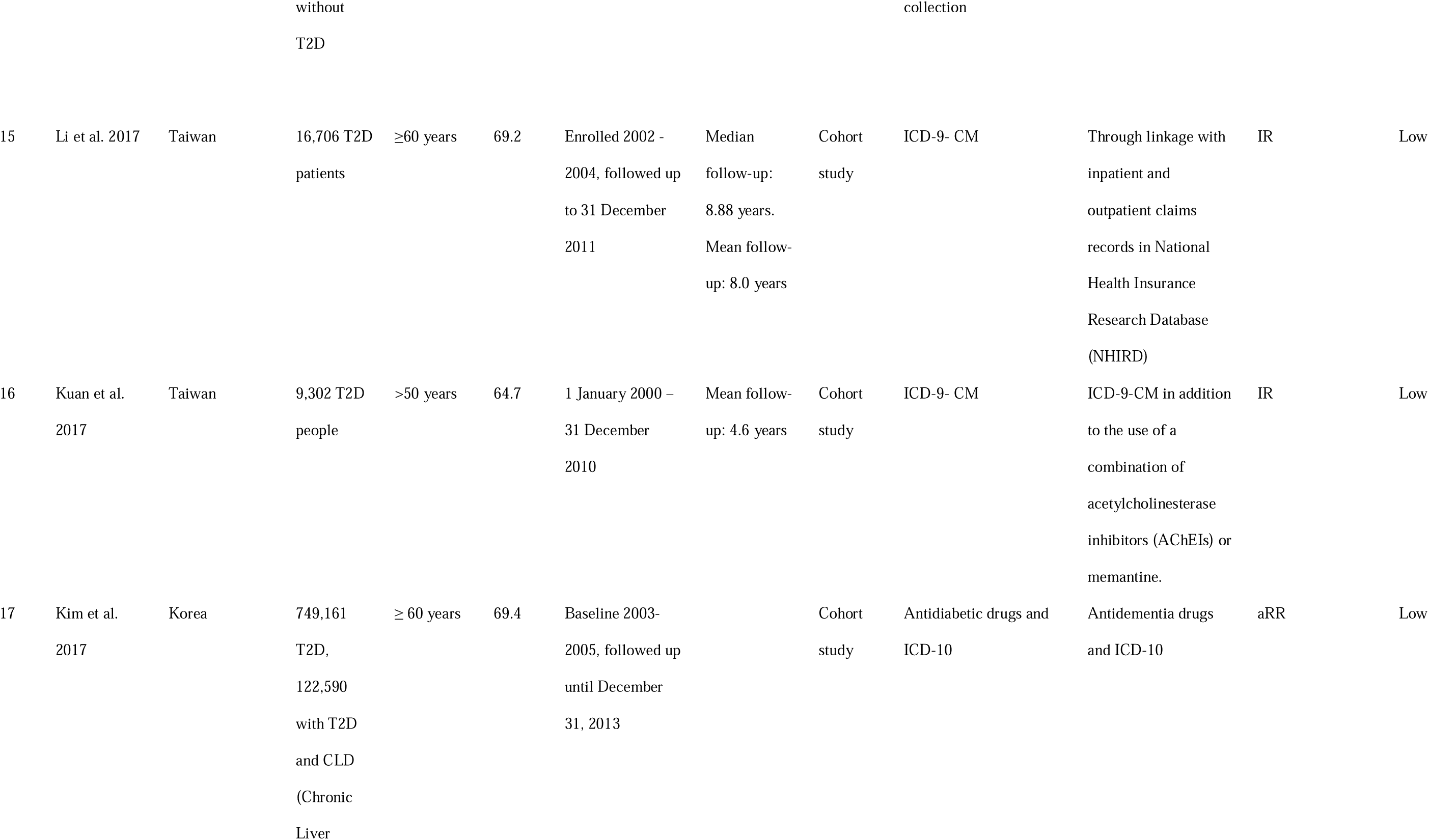
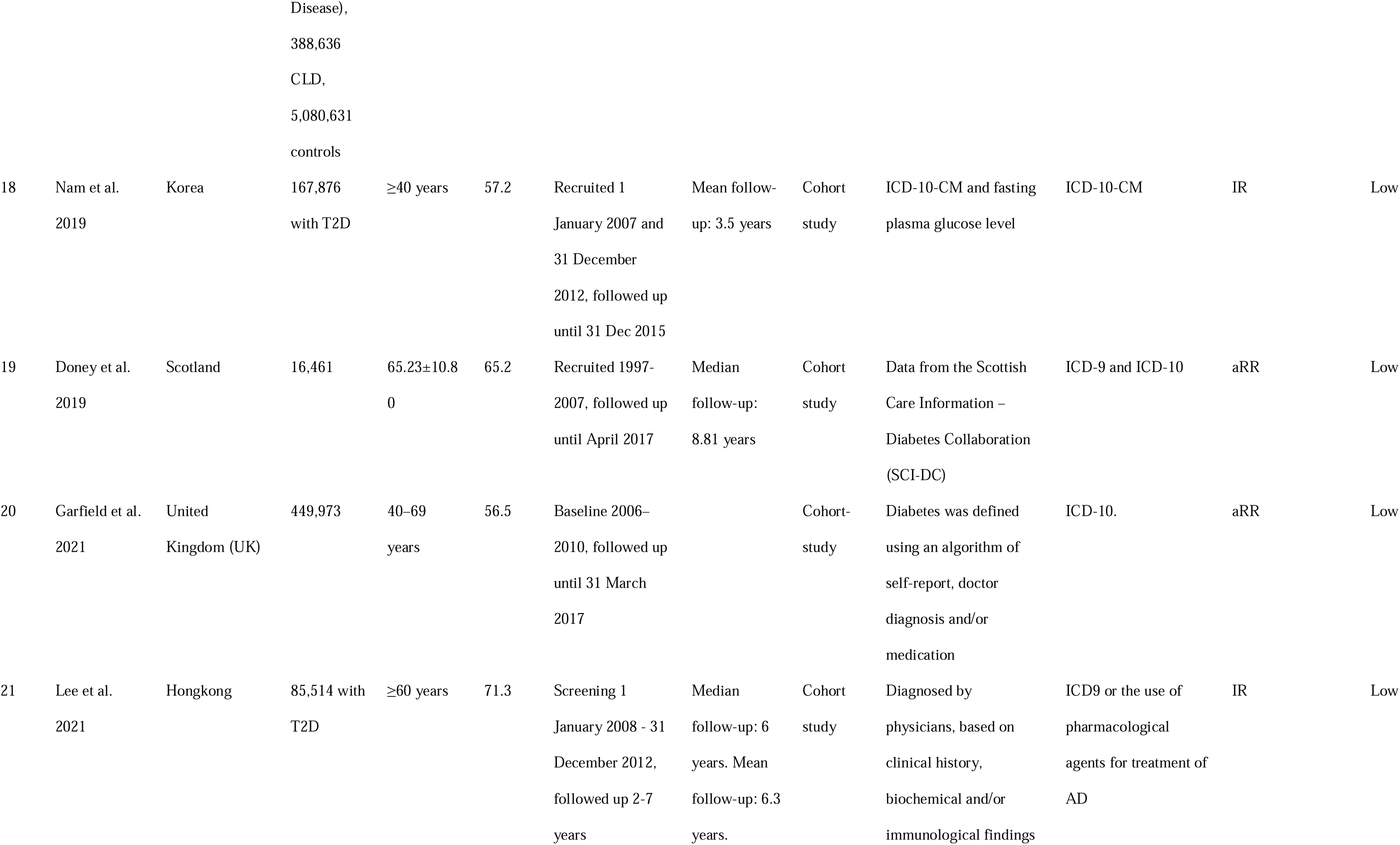
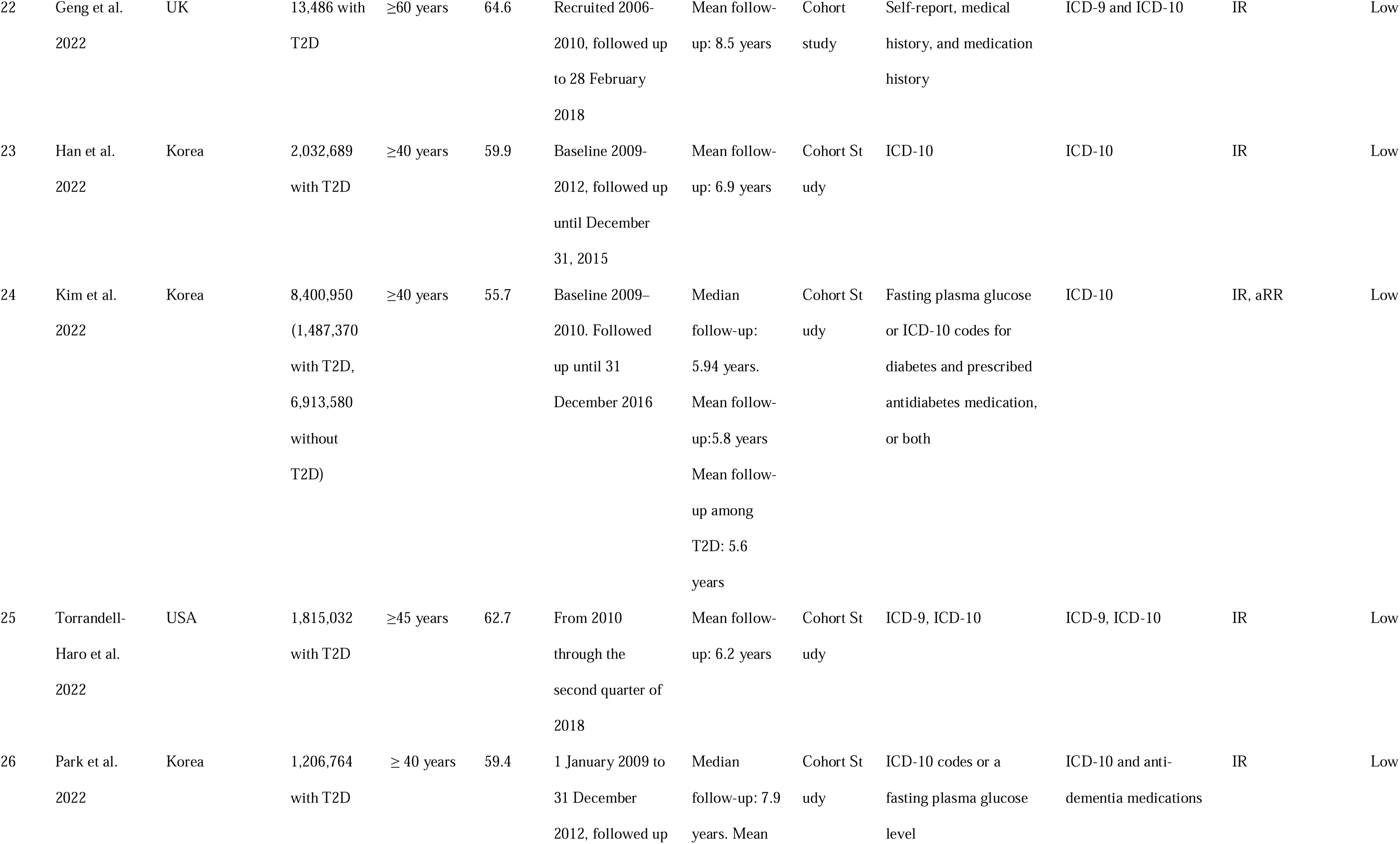
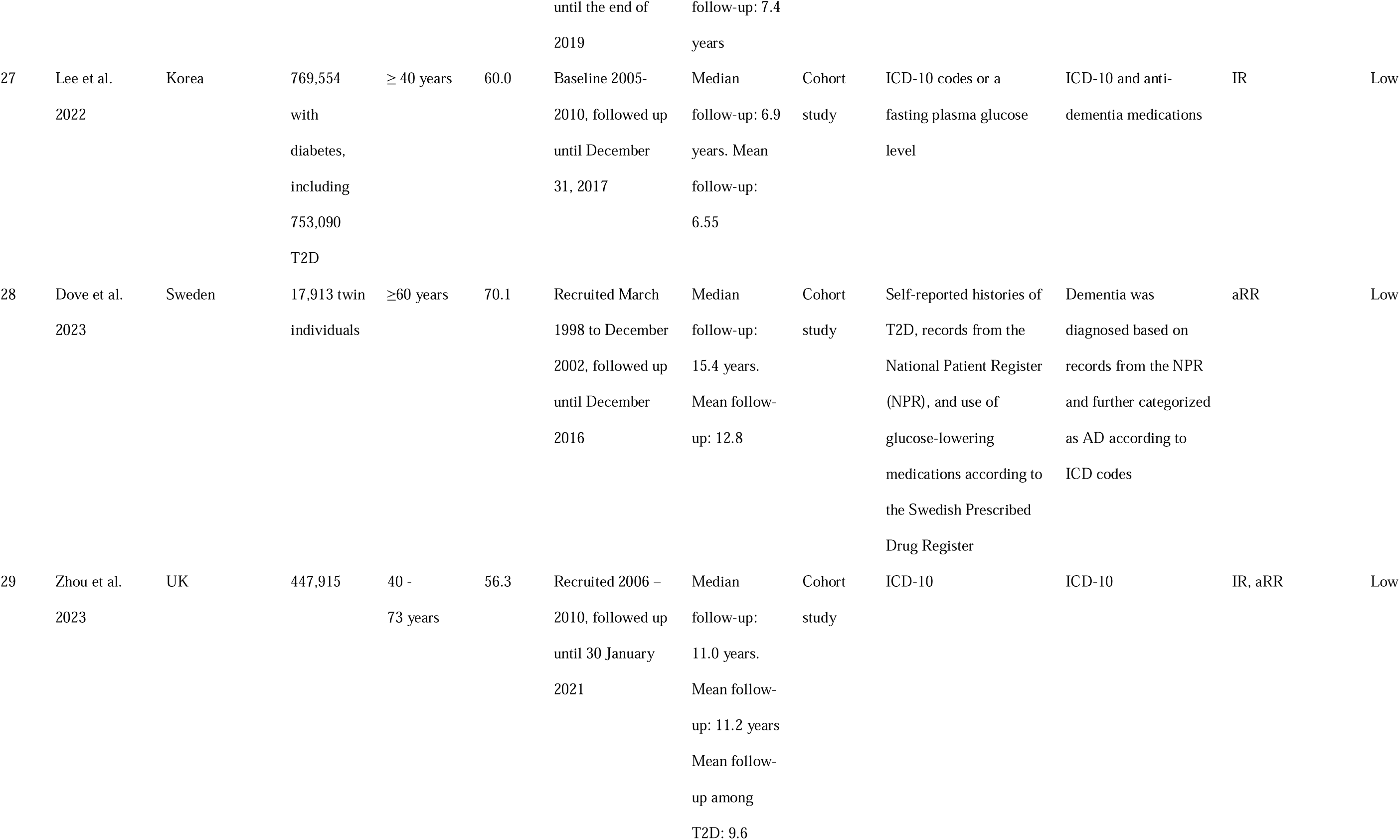
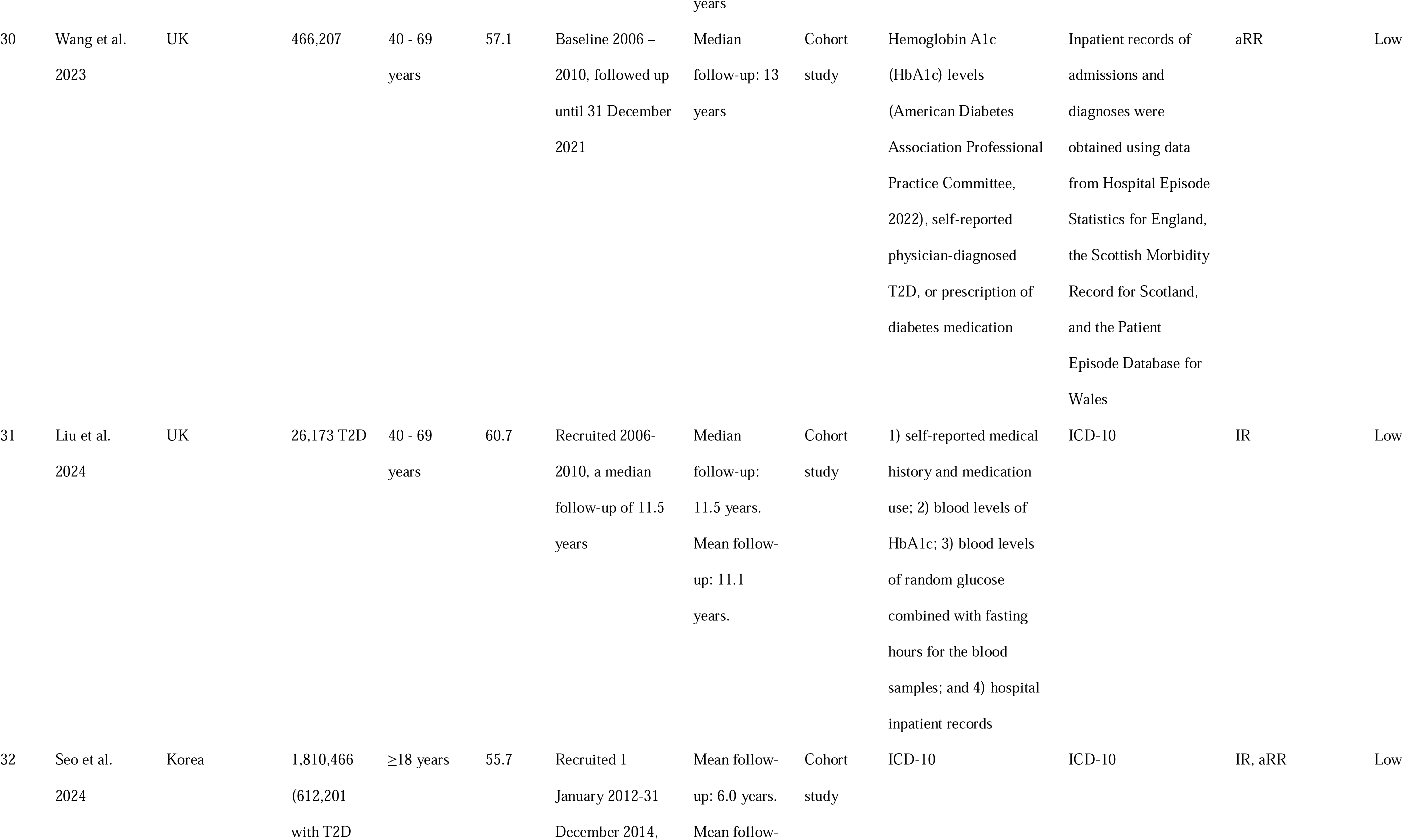
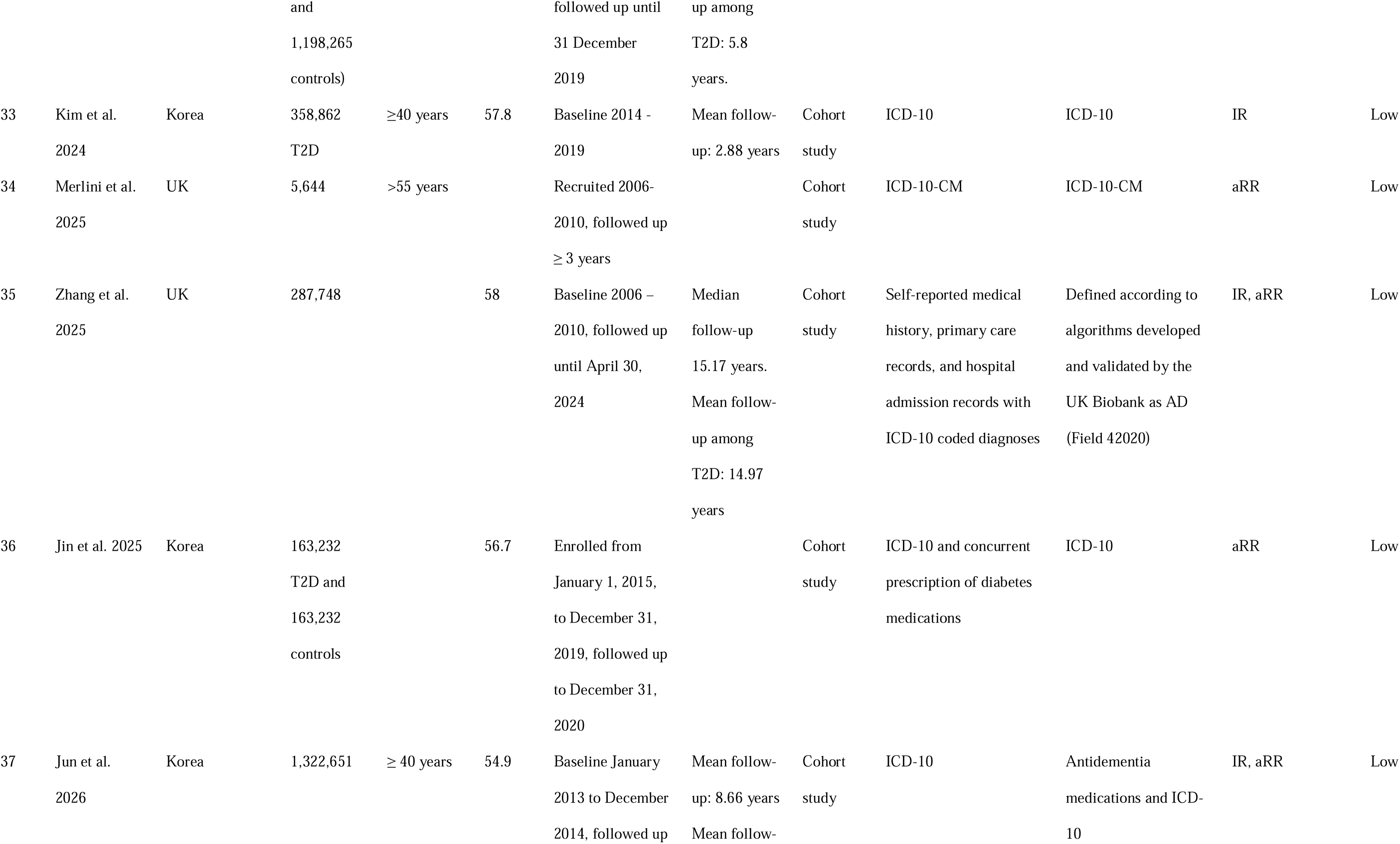
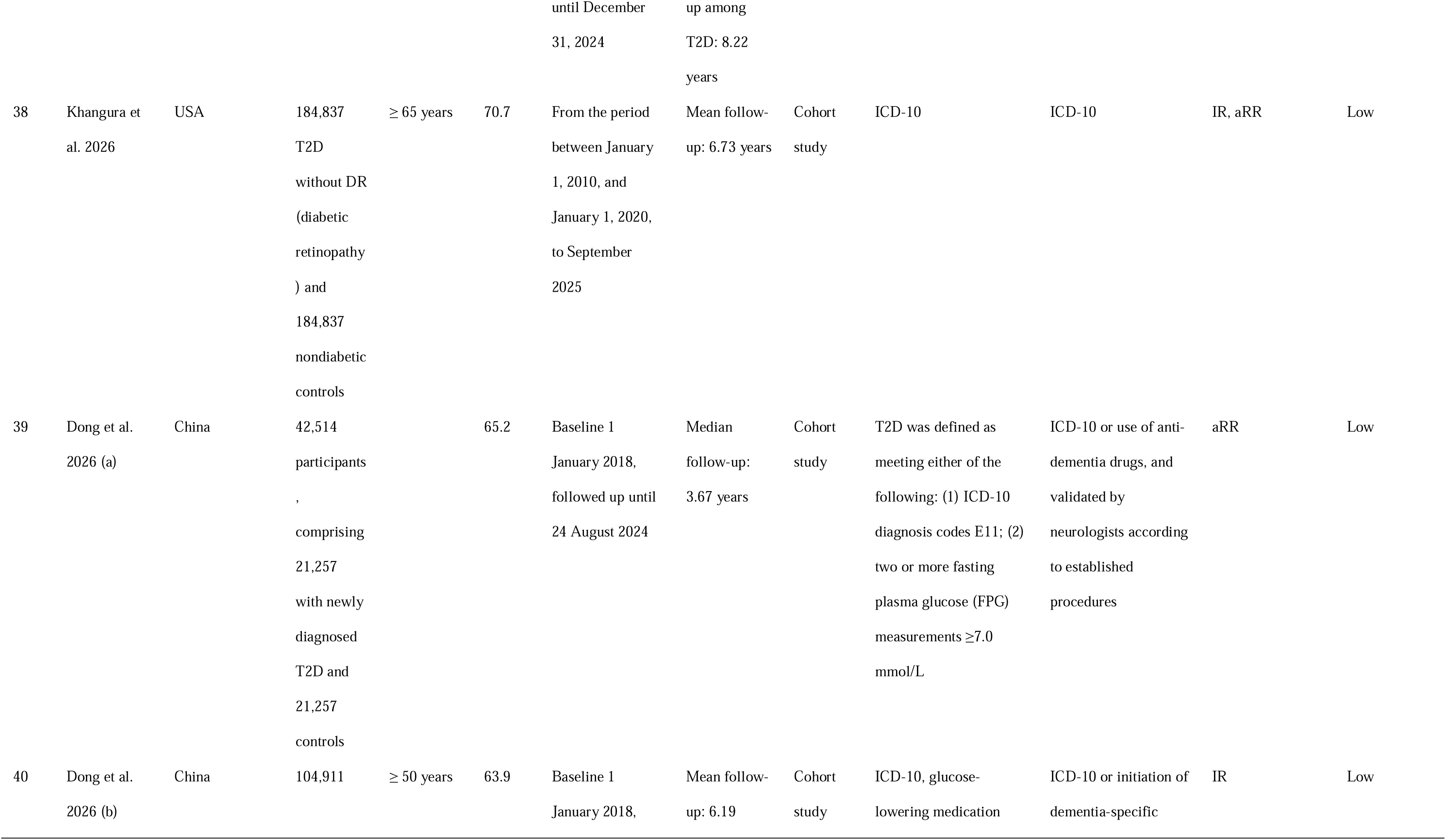
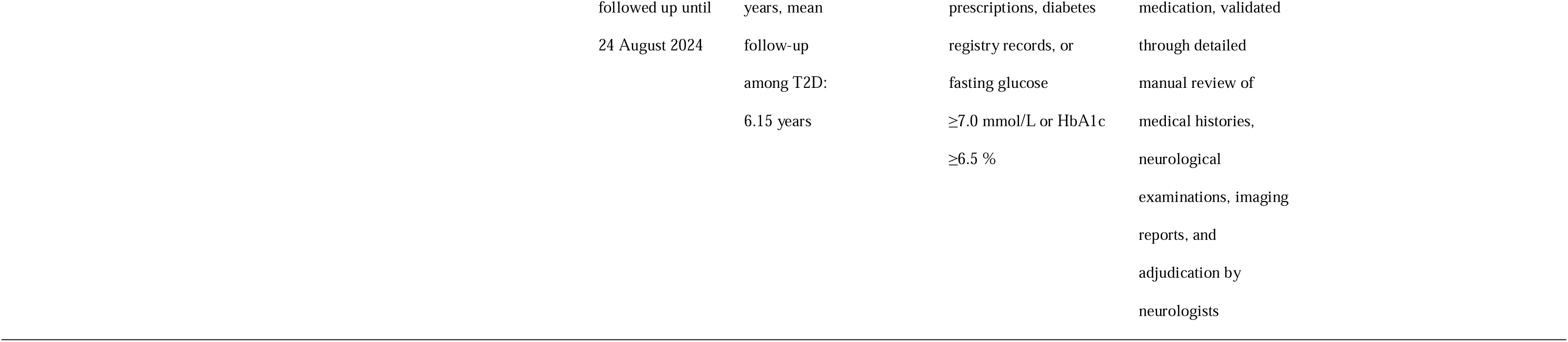
Characteristics of the included studies.

### Eligibility criteria and selection process

This systematic review included observational studies published in peer-reviewed journals that investigated the incidence of AD among individuals with T2D or assessed the association between T2D and AD. Studies were eligible if they reported incidence rates (IRs), adjusted effect estimates including odds ratios (aORs), relative risks (aRRs), or hazard ratios (aHRs), or provided sufficient data to allow these measures to be calculated.

The following studies were excluded:

- Studies that did not provide disaggregated data specific to T2D and AD, including those focusing on other diabetes subtypes (type 1 diabetes or gestational diabetes) or non-AD dementias (e.g., vascular or Lewy body dementia)
- Clinical trials, Mendelian randomisation studies, and review articles
- Case reports, editorials, letters, and conference abstracts lacking sufficient data
- Animal or in vitro studies
- Duplicate publications derived from the same dataset

The search results were imported into a citation management software (EndNote version 21.4), and duplicate records were removed, with only the most comprehensive or most recent version retained. All remaining references were then uploaded to Rayyan [30], an online platform designed to facilitate screening for systematic reviews, where four (TTHN, CIO, AA and EOA) investigators independently screened titles and abstracts to identify eligible studies. These were then further screened against inclusion and exclusion criteria, with any discrepancies resolved through consensus.

### Data extraction

Data from each eligible study were extracted using a piloted template designed in Microsoft® Excel® for Microsoft 365 MSO (Microsoft Office Suite) Version 2308. The data were extracted by one investigator (TTHN) and independently checked by two other investigators (EAD and AA) acting as second reviewers. Any discrepancy in data extraction was resolved by consensus. The extracted data included the authors, year of publication, study location and design, sample size, participant demographic characteristics, follow-up duration, diagnostic criteria for T2D and AD, incidence of AD, and outcome measures of association, including aORs, aRRs, or aHRs with corresponding 95% confidence intervals (CIs). Only multivariable-adjusted estimates were extracted. From studies reporting multiple multivariable models, we extracted estimates from those with the most comprehensive adjustment set, as fully adjusted models better account for potential confounding and provide the most reliable estimates of the independent association between T2D and AD.

### Study risk of bias assessment

Cohort and case-control studies were assessed for risk of bias using the Newcastle–Ottawa Scale (NOS) [31]. To assess the risk of bias of cross-sectional studies, an adapted version of the NOS scale was used [32]. All the risk-of-bias tools evaluated studies across three domains (selection of study groups, comparability of groups, and ascertainment of the exposure or outcome), with a maximum possible score of 9 stars. Total stars of 7-9 were interpreted as indicative of low risk of bias, stars of 4-6 as moderate risk, and stars of 0-3 as high risk. Risk-of-bias assessments were conducted by one investigator (TTHN) and independently verified by two additional investigators (EAD and AA).

### Data analysis

Statistical analyses were performed using Stata version 19.5 (StataCorp. LLC, College Station, USA). Pooled estimates were derived using a random-effects meta-analysis, with between-study variance estimated using the Restricted Maximum Likelihood (REML) method. Data on the IRs per 1,000 person-years (PY) of AD in patients with T2D were pooled to derive an overall pooled estimate. A 95% prediction interval (PI) was calculated for the pooled IR to capture between-study variability, using the formula PI = pooled estimate ± t-critical × √(SE² + τ²), where t-critical was derived from a t-distribution with k − 2 degrees of freedom (k = number of studies included in the meta-analysis). Adjusted relative risks (aRRs) were used as the standard measure of association across the included studies. Odds ratios (ORs) were converted to RRs using the formula: RR = OR / [(1 − P=) + (P= × OR)], where P= denotes the prevalence of the outcome in the non-exposed (control) group [33]. Hazard ratios (HRs) were treated as equivalent to relative risks (RRs). As this assumption may be less valid in older populations with higher cumulative event rates, we conducted a subgroup analysis comparing pooled estimates from studies that contributed aRRs derived from aHRs versus those that contributed aRRs derived from aRRs or aORs. Additionally, a leave-one-out sensitivity analysis was conducted among cohort studies to assess the influence of individual studies on the pooled aRR estimate.

Interstudy heterogeneity was assessed using Cochran’s Q test, which provides a chi-square (χ2) statistic and corresponding P-value. The percentage of the total variation across studies attributable to heterogeneity was estimated using Higgins and Thompson I^2^ statistic [34]. Sources of heterogeneity were explored through subgroup analyses for both IR and aRR analyses. Additionally, univariable meta-regression was conducted for continuous covariates only, as subgroup analysis of categorical covariates provides comparable results to meta-regression with categorical predictors. Publication bias or small-study effect was assessed using funnel plots and Egger’s regression test [35].

Subgroup analyses were conducted separately for IRs per 1,000 PY and aRRs. For both analyses, studies were stratified by publication year, geographic region, mean or median age, mean or median follow-up duration, and methods used to assess T2D and AD. For the aRR analysis only, stratification additionally included the number of adjusted confounders and the outcome measure used to derive the aRR (studies reporting aHR versus those reporting aRR or aOR). Geographic regions were classified according to the World Bank regional classification [36]. Pooled estimates were calculated within subgroups using a random-effects meta-analysis, and differences between subgroups were assessed using the between-group Cochran’s Q test.

## Results

### Study selection

A total of 9,430 records were identified through database searches, of which 532 duplicates were removed, leaving 8,898 records for eligibility screening. One additional record was identified through citation searching and included directly. After screening and eligibility assessment, 40 studies [37–76] covering 27,102,559 participants were included in the meta-analyses (Figure 1).

**Figure 1.**
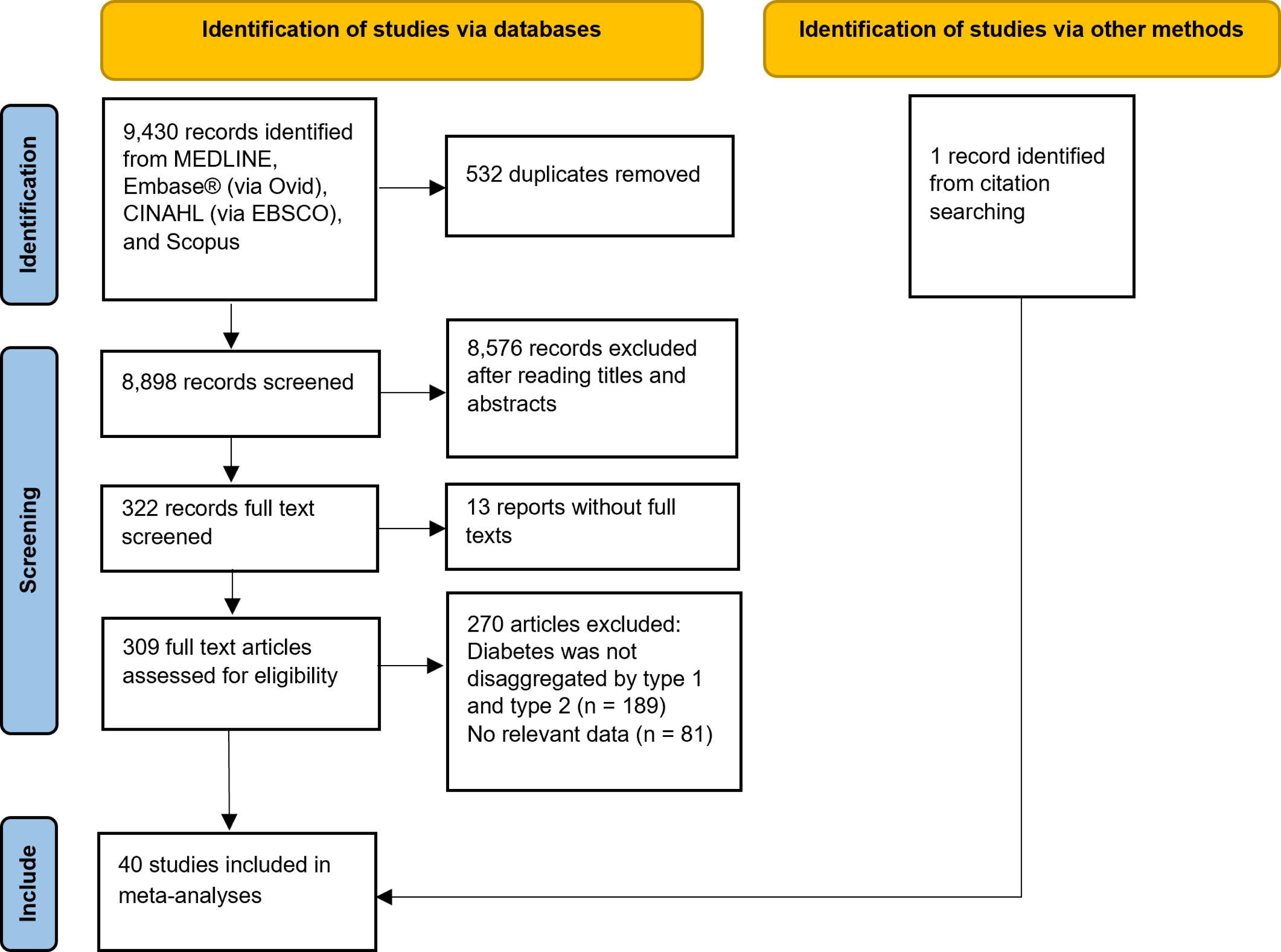
Flow diagram of study selection process.

### Characteristics of the included studies

The main characteristics of the included studies are presented in Table 1. The studies were published between 1997 and 2026 and varied widely in sample size, ranging from 702 to 8,400,950 participants. Most studies employed a cohort design (38/40), with only one case–control study and one cross-sectional study. Among cohort studies, mean or median follow-up duration ranged from 2.0 to 15.2 years. The included studies were conducted across 12 countries, with the largest number originating from Korea (n=10), the United Kingdom (UK) (n=7), the United States (US) (n=8), and Taiwan (n=4), followed by Sweden (n=3), China (n=2), and one study each from the Netherlands, France, Egypt, Australia, Scotland, and Hong Kong. Ten Korean studies drew on national health insurance or administrative databases, seven UK studies utilised the UK Biobank, three Taiwanese studies used the Taiwan National Health Insurance Research Database (NHIRD), and two US studies used large administrative claims databases. The remaining studies were based on dedicated population-based cohort datasets. The age range varied across studies, with most focusing on older or middle-aged populations. The most common minimum age criteria were ≥65 years (n=7), ≥40 years (n=7), and ≥60 years (n=5). The mean or median ages, as reported in individual studies, ranged from 55 to 75 years in most studies.

#### Assessment of type 2 diabetes

Twenty-five studies used administrative data to ascertain T2D, primarily through ICD (International Classification of Diseases) codes (version 7 through 10) drawn from hospital records, national registries, or insurance claims databases. These were often supplemented by additional methods, including prescription records for diabetes related medications, fasting plasma glucose levels, or HbA1c measurements. The remaining 15 studies relied exclusively on non-administrative methods, which varied across studies and included self-reported physician diagnosis, structured questionnaires, clinical examination, laboratory-based criteria such as fasting plasma glucose or HbA1c, and medication use.

#### Assessment of Alzheimer’s disease

AD was ascertained using administrative methods in 28 studies, most commonly through ICD codes from hospital or national health records, frequently supplemented by other methods such as prescription records for anti-dementia medications, or further validated by neurologist review. The remaining 12 studies relied exclusively on non-administrative criteria, which varied across studies and included the NINCDS-ADRDA (National Institute of Neurological and Communicative Disorders and Stroke–Alzheimer’s Disease and Related Disorders Association) criteria, the Diagnostic and Statistical Manual of Mental Disorders (DSM-III-R), and the Mini-Mental State Examination.

#### Risk of bias

Most studies (38/40) were classified as low risk of bias, reflecting robust designs, large sample sizes, and reliable exposure and outcome ascertainment. Two studies were rated as moderate risk, primarily due to limitations in exposure assessment (Table 2S – Supplementary materials). No studies were classified as high risk of bias.

### Incidence rates analysis

#### Meta-analysis of incidence rates of Alzheimer’s disease among individuals with type 2 diabetes

Of the 40 studies included in this review, 23 contributed data suitable for calculating the IR of AD in individuals with T2D per 1,000 PY. These studies included 8,961,034 participants and had a cumulative follow-up of 56,484,094 PY. Reported IRs ranged from 0.88 per 1,000 PY to 13.91 per 1,000 PY. The pooled random-effects IR of AD in individuals with T2D was 4.71 per 1,000 PY (95% CI 3.31–6.71) (Figure 2). Heterogeneity across studies was extremely high (I² = 99.99%), indicating substantial variability in incidence estimates among the included studies. The 95% prediction interval (0.76 to 29.30 per 1,000 PY) captures the extent of between-study variability, indicating that the true IR of AD in individuals with T2D could plausibly range from less than 1 to nearly 30 per 1,000 PY in a new study drawn from the same distribution. Egger’s regression test did not demonstrate significant small-study effects (β= = −0.07, SE = 2.561, z = −0.03, *P-value* = 0.977). Visual inspection of the funnel plot (Figure 1S-Supplementary materials) suggested minor asymmetry; however, the non-significant Egger’s test indicates that this is unlikely due to small-study effects or publication bias.

**Figure 2.**
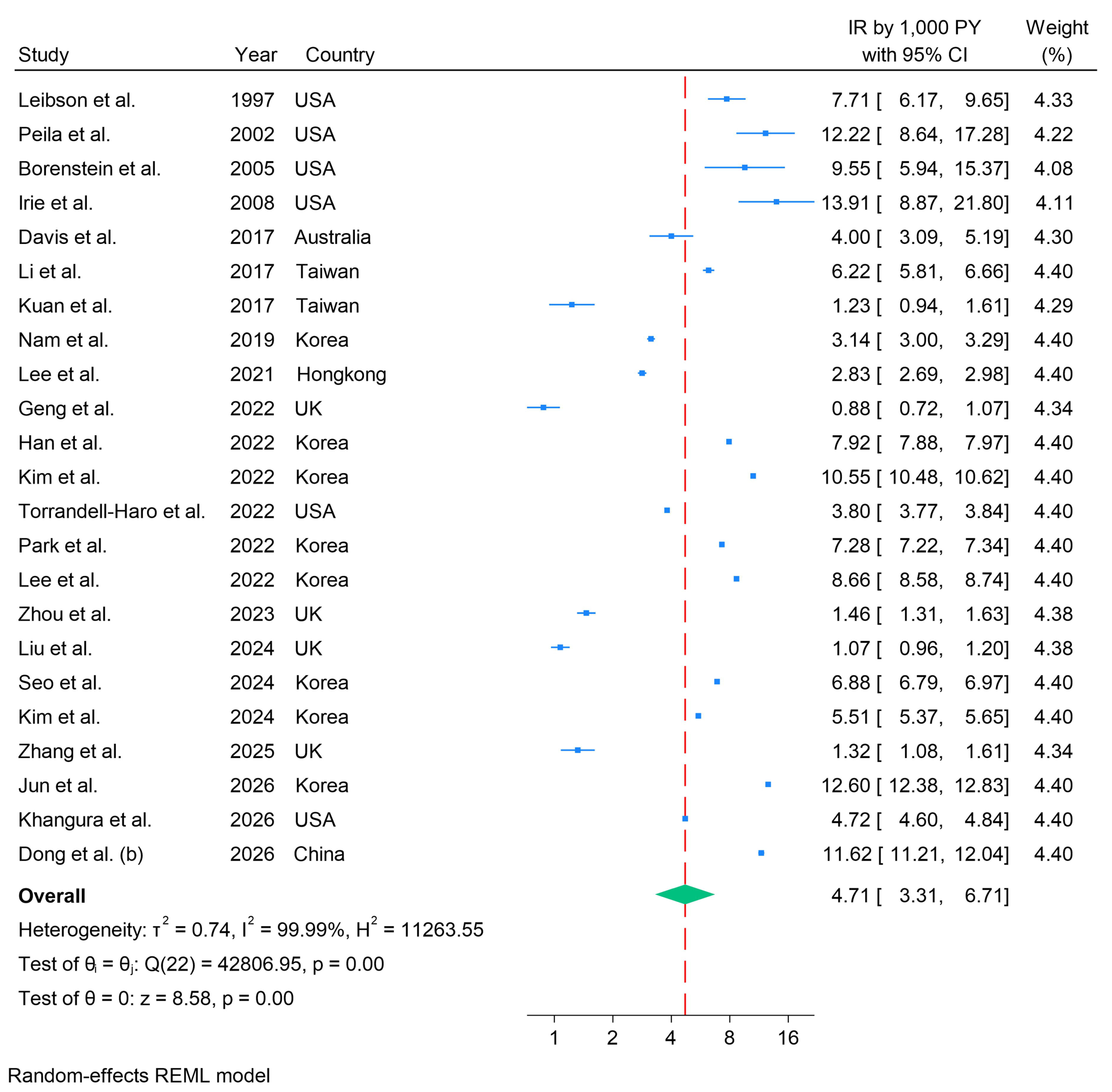
Meta-analysis of incidence rates of Alzheimer’s disease among individuals with type 2 diabetes.

#### Meta-regression analyses of incidence rates of Alzheimer’s disease among individuals with type 2 diabetes: continuous covariates

Mean follow-up duration was a significant predictor of heterogeneity in meta-regression (coefficient = −0.129, 95% CI: −0.240 to −0.019, p = 0.022), explaining 16.37% of between-study variance; longer follow-up was associated with lower IR of AD. Publication year (p = 0.131) and mean age (p = 0.416) were not significant explanatory variables for heterogeneity,

#### Subgroup analyses of incidence rates of Alzheimer’s disease among individuals with type 2 diabetes

IRs did not significantly differ by publication year (*P-value* = 0.624), mean/median age (*P-value* = 0.743), mean follow-up duration (*P-value* = 0.406), or T2D ascertainment method (*P-value* = 0.709) (Figure 3). However, significant differences were observed by region (*P-value* < 0.001) and AD ascertainment method (*P-value* <0.001). The pooled IR was highest in North America (7.54 per 1,000 PY, 95% CI 4.94–11.51), followed by East Asia and the Pacific (5.81 per 1,000 PY, 95% CI 4.08–8.28), and lowest in Europe and Central Asia (1.17 per 1,000 PY, 95% CI 0.94–1.45). Studies using administrative methods (alone or combined with additional methods) to assess AD reported substantially lower incidence rates (4.00 per 1,000 PY, 95% CI 2.72–5.89) than those using non-administrative methods (10.24 per 1,000 PY, 95% CI 7.71–13.60).

**Figure 3.**
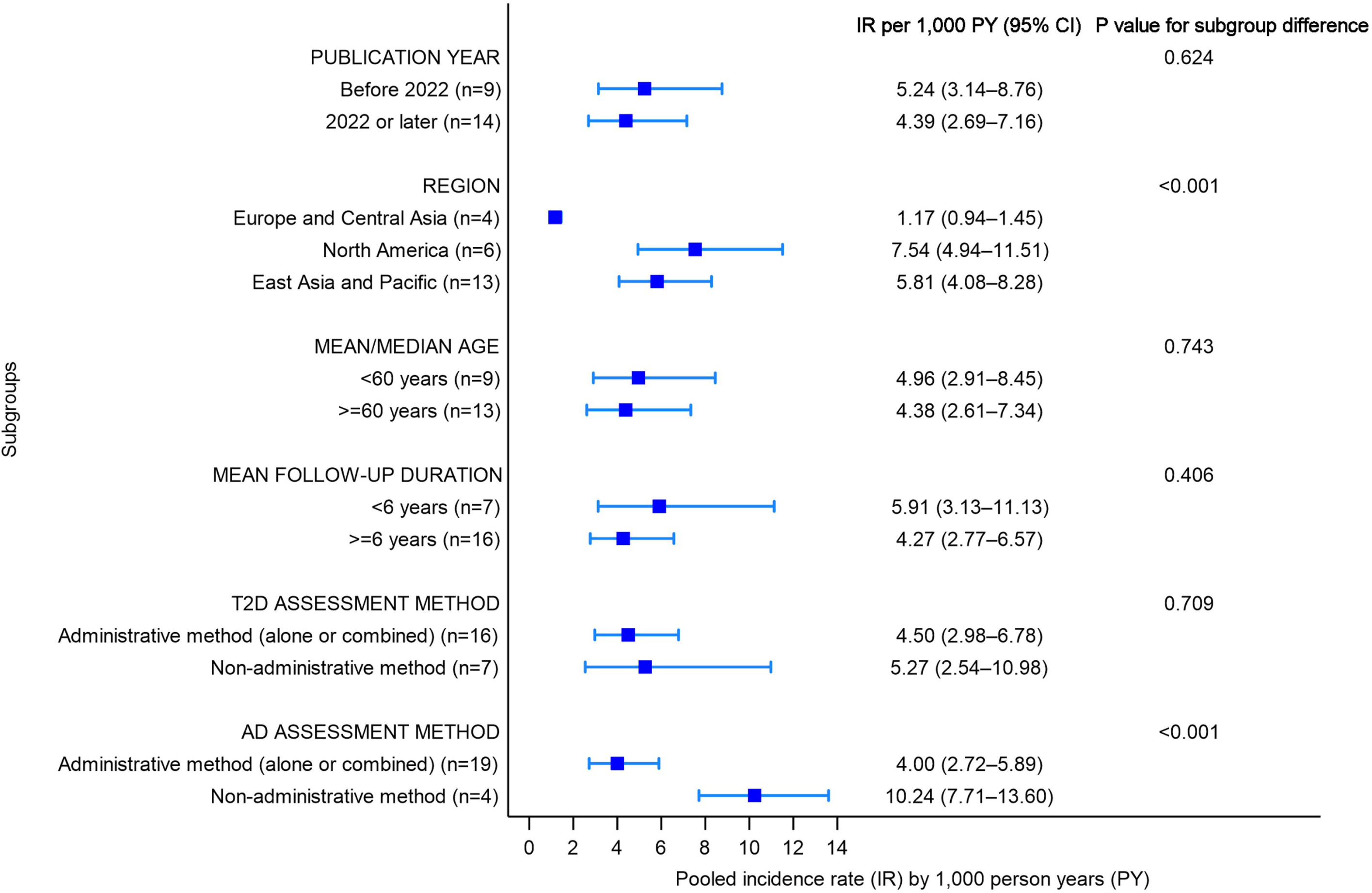
Subgroup analyses of incidence rates of Alzheimer’s disease among individuals with type 2 diabetes

### Adjusted relative risks analysis

#### Meta-analysis of adjusted relative risks of Alzheimer’s disease among individuals with type 2 diabetes

Among the 40 included studies, 26 contributed data on the association between T2D and AD as aRRs, comprising aRRs derived from aHRs, aRRs, or aORs. Across studies, aRRs were derived from multivariable models, most consistently controlling for age and sex, with many also adjusting for educational level; other covariates include ethnicity, family history, marital status, region, body mass index (BMI), lifestyles (smoking, alcohol consumption, vegetarian diet, physical activities, and leisure activities), *APOE*-ε4 status, medications, and comorbidities. The pooled aRR was 1.53 (95% CI 1.38–1.70) (Figure 4), indicating a statistically significant increased risk of AD after accounting for potential confounding factors. Heterogeneity remained extremely high (I² = 99.60%), indicating substantial variability in effect estimates across studies. Egger’s regression test did not indicate the presence of small-study effects (β= = 0.08, SE = 0.621, z = 0.13, *P-value* = 0.895). Consistent with this finding, the funnel plot (Figure 2S – Supplementary materials) did not show an extremely asymmetric distribution of studies around the pooled effect estimate. Assessment of small-study effects did not reveal evidence of publication bias. Leave-one-out sensitivity analysis showed that the pooled aRR remained stable across all iterations (range: 1.50 to 1.57), with all estimates remaining statistically significant (all p < 0.001), indicating that no single study disproportionately influenced the overall finding.

**Figure 4.**
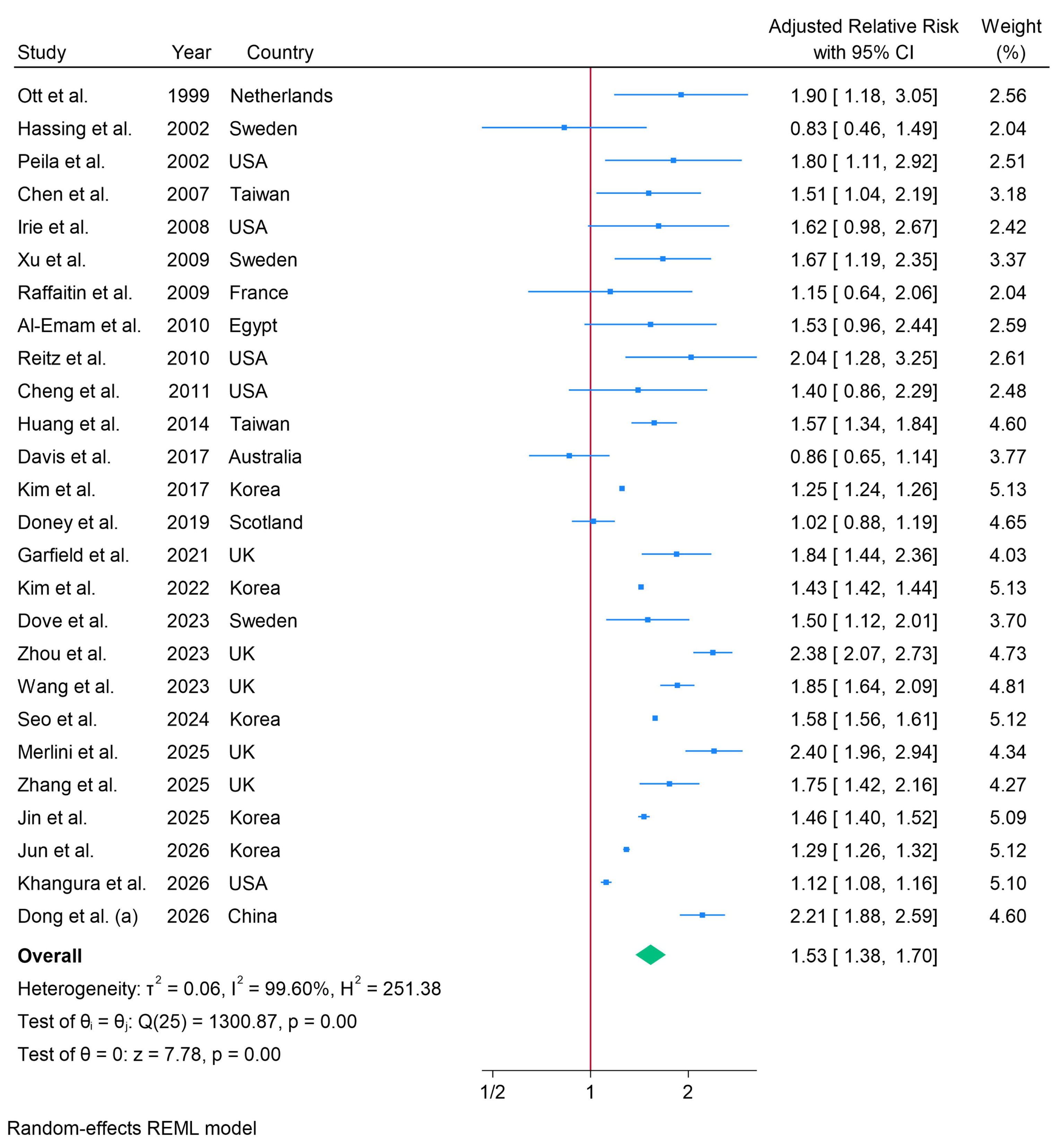
Meta-analysis of adjusted relative risks of Alzheimer’s disease among individuals with type 2 diabetes

#### Meta-regression analyses of adjusted relative risks of Alzheimer’s disease among individuals with type 2 diabetes: continuous covariates

Publication year (p = 0.524), mean/median age (p = 0.240), and mean/median follow-up duration (p = 0.589) were not significantly associated with between-study heterogeneity.

#### Subgroup analysis of adjusted relative risks of Alzheimer’s disease among individuals with type 2 diabetes

Subgroup analyses of aRRs showed consistent positive associations across publication period, geographic region, mean/median age, mean/median follow-up duration, and methods used to ascertain T2D or AD. Pooled estimates ranged from 1.40 to 1.66 across subgroups (Figure 5). None of the subgroup differences was statistically significant (all *P-values* for subgroup differences >0.05).

**Figure 5.**
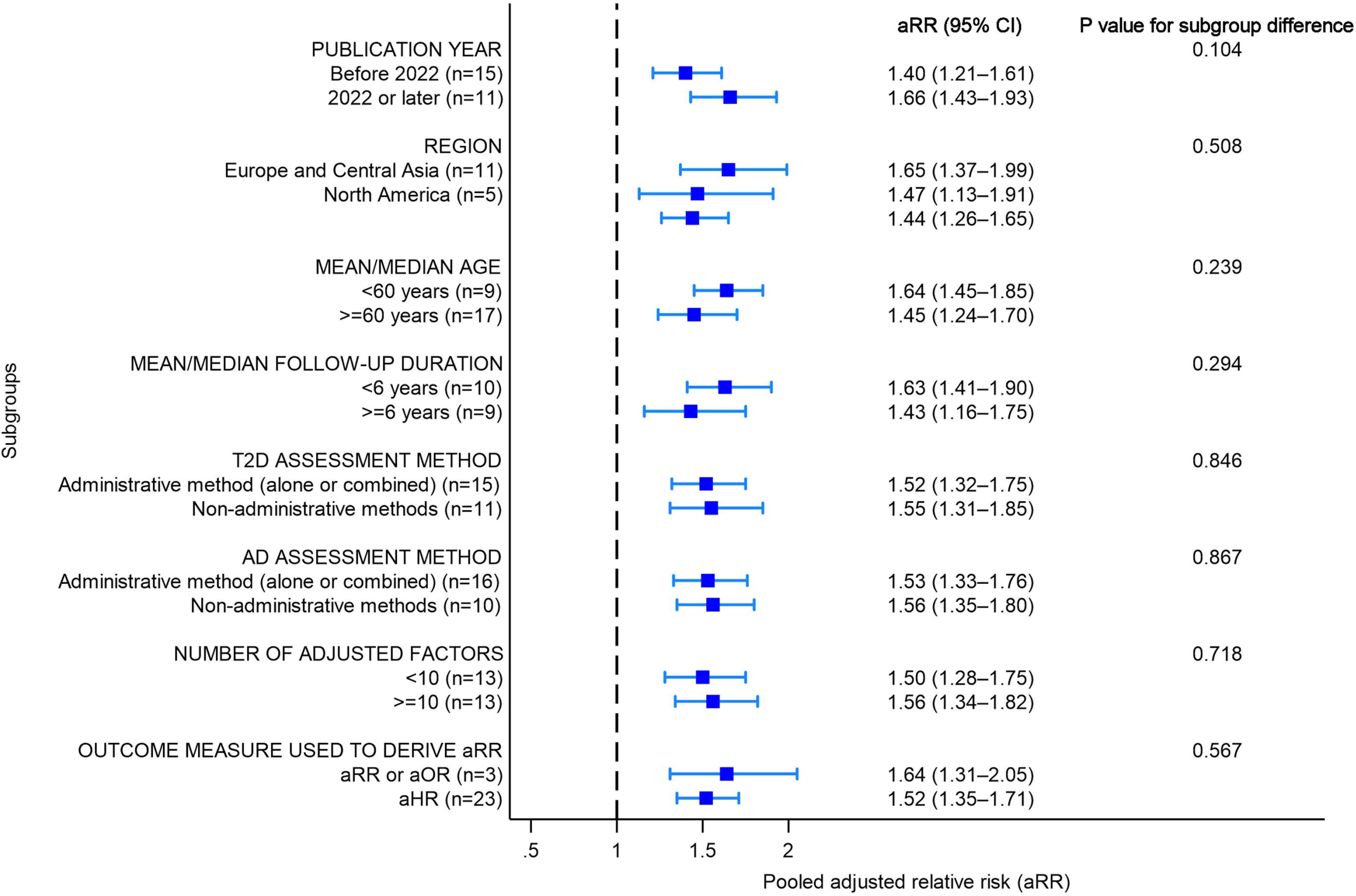
Subgroup analyses of adjusted relative risks of Alzheimer’s disease among individuals with type 2 diabetes

## Discussions

In this systematic review and meta-analysis of 40 observational studies, including 27,102,559 participants, we found that T2D was associated with a significantly increased risk of AD. Across adjusted analyses, individuals with T2D had a 53% higher relative risk of AD, and the pooled incidence rate among people with T2D was 4.71 cases per 1,000 person-years, providing a clinically interpretable estimate of absolute disease burden. Importantly, the association remained directionally consistent across subgroup analyses stratified by geography, age, follow-up duration, and exposure and outcome ascertainment methods, despite substantial between-study heterogeneity.

One of the most clinically informative findings of this study is the pooled incidence estimate of 4.71 AD cases per 1,000 person-years among individuals with T2D. While relative risks are valuable for etiological inference, incidence rates provide a direct measure of disease burden that is highly relevant for risk stratification, prevention planning, and health-system forecasting [77, 78]. The higher incidence observed in North America compared with Europe and Australia may reflect regional differences in cardiometabolic risk profiles, health-care access, and case ascertainment, although these potential explanations require direct empirical evaluation [79]. These findings support the value of context-specific dementia prevention approaches and underscore the importance of comprehensive management of modifiable risk factors in people with T2D [80]

These findings reinforce and extend prior epidemiological evidence linking diabetes with neurodegenerative outcomes [11, 13, 14, 16]. Previous meta-analyses consistently reported an elevated risk of dementia among individuals with diabetes; however, most pooled all-cause dementia, vascular dementia, and AD together, or combined heterogeneous diabetes phenotypes [11, 14, 16]. Such approaches may risk blending mechanistically distinct disease pathways, particularly given the strong associations between diabetes, cerebrovascular disease, and mixed dementia pathology [22–24]. By restricting inclusion to studies with well-defined T2D exposures and AD outcomes, our synthesis addresses a major limitation of earlier reviews of conflating diabetes subtypes and broad dementia outcomes and provides a more biologically coherent estimate of AD-specific risk.

The biological plausibility of the association found in our study is well supported in the literature. Insulin resistance, chronic hyperglycaemia, mitochondrial dysfunction, oxidative stress, and neuroinflammation may jointly promote amyloidogenic processing, tau hyperphosphorylation, synaptic dysfunction, and neuronal loss [17–19, 21]. Recent molecular-level syntheses further implicate shared dysregulation across insulin signalling, lipid metabolism, inflammatory cascades, and epigenetic pathways that link T2D to late-onset AD [81]. However, recent genetic studies suggest that liability to T2D alone is unlikely to be sufficient to cause AD, with cross-omic analyses indicating that the relationship does not conform to a simple genetic shared-risk architecture [82, 83]. Instead, the association may arise from complex genetic variation acting across molecular pathways, with opposing effects on disease risk [83]. These findings suggest that the observed association is driven mainly by metabolic, vascular, or treatment-related factors rather than inherited genetic risk. This broader evidence may help reconcile the consistent observational association observed here with comparatively weaker causal or shared genetic signals reported elsewhere[82, 83].

The very high heterogeneity observed across pooled analyses warrants careful interpretation. Rather than undermining the association, this heterogeneity likely reflects the real-world complexity of both conditions, as illustrated by the wide prediction interval for the pooled incidence rate (0.76 to 29.30 per 1,000 PY), indicating that true AD incidence rates in individuals with T2D may vary substantially across different populations and settings. T2D encompasses a wide variation in disease duration, severity, glycaemic control, insulin resistance, obesity profiles, medication exposure, and complication burden [84]. Similarly, AD diagnosis varies substantially according to whether ascertainment is based on specialist clinical criteria, neuropsychological assessment, medication proxies, or administrative coding [85]. Our subgroup analyses are informative in this regard: studies using non-administrative AD ascertainment methods reported substantially higher incidence rates than registry-based studies, suggesting that diagnostic sensitivity and outcome ascertainment materially influence absolute risk estimates. This observation likely reflects under-ascertainment of milder AD cases in administrative data and more rigorous clinical adjudication in research-based cohorts.

The principal novelty of this study lies in its restriction to well-defined T2D and AD populations and its comprehensive synthesis of incidence and relative risk estimates. To our knowledge, no previous meta-analysis has simultaneously (i) restricted inclusion to clearly defined T2D and AD phenotypes, and (ii) separately synthesised and analysed incidence rates alongside adjusted relative risk estimates. By excluding studies with mixed diabetes phenotypes and non-specific dementia outcomes, and by applying consistent selection criteria, we present the relationship between T2D and AD with greater disease specificity than prior syntheses. Furthermore, by integrating incidence estimates with relative risk metrics, our analysis provides a clinically interpretable perspective that remains uncommon in prior reviews.

Our findings also have implications for the emerging literature on diabetes therapeutics and dementia prevention. Observational and network meta-analytic evidence suggest that some glucose-lowering therapies, particularly GLP-1 receptor agonists and SGLT2 inhibitors, may be associated with lower dementia risk among individuals with T2D [86]. Although causal inference is limited, these estimates provide a quantitative benchmark for future target-trial emulations investigating whether metabolic interventions can alter AD risk.

Several limitations are acknowledged. First, all included studies were observational, precluding causal inference and leaving residual confounding possible. Second, although meta-regression and subgroup analyses identified meaningful sources of heterogeneity in incidence rates, particularly by follow-up duration, region, and AD ascertainment method, substantial heterogeneity persisted overall. This residual variability was especially evident in relative risk estimates and likely reflects unmeasured differences in T2D duration and severity, glycaemic control, medication exposure, *APOE* genotype, and competing mortality risks, which were inconsistently reported. In addition, subgroup analyses were constrained by the availability and consistency of reported data and should be interpreted cautiously.

Third, although we prioritised phenotypic specificity, AD diagnosis in several large population-based studies relied on ICD codes or medication proxies, raising the possibility of outcome misclassification. Fourth, the included studies spanned multiple decades and were conducted largely in high-income settings, which may limit generalisability to other populations and reflect temporal changes in diagnostic criteria and clinical management of both T2D and AD. Fifth, definitions of diabetes exposure varied across studies, with some examining glycaemic variability, medication use, or cardiometabolic multimorbidity rather than T2D as a binary exposure, potentially introducing heterogeneity in effect estimates. Sixth, we did not synthesise prevalence estimates, because they reflect both disease occurrence and survival. Instead, we focused on estimates of incidence and relative risk, which more directly characterise the association between T2D and AD. Finally, although a subgroup analysis of aRRs comparing studies with < 6 years versus ≥ 6 years of follow-up demonstrated consistent effect estimates, reverse causation cannot be entirely excluded.

Future individual patient-level meta-analysis would enable more precise confounding control, better address reverse causation, and provide more accurate AD incidence estimates among individuals with T2D.

### Conclusions

Our meta-analysis provides robust epidemiological evidence that T2D is associated with a significantly increased risk of AD, even when both conditions are stringently defined. By prioritising disease specificity, harmonised methodology, and clinically interpretable incidence estimates, this study advances the field beyond earlier broad dementia syntheses and clarifies the magnitude of the T2D–AD association as an AD-specific outcome. These findings support integrating cognitive risk surveillance into long-term diabetes care and underscore the need for mechanistically informed studies examining how glycaemic trajectories, metabolic phenotypes, and contemporary glucose-lowering therapies may influence AD pathogenesis.

## Declarations

### Ethics approval and consent to participate

Not applicable

### Consent for publication

Not applicable

### Availability of data and materials

All data generated or analysed during this study are included in this published article and its supplementary information files

### Authors’ contributions

Conceptualisation and design: AA and EOA. Literature search and retrieval: AA and EOA. Article screening: TTHN, EOA, AA, CIO, and VO. Data extraction: TTHN, AA, EOA, and EAD. Data analysis: TTHN, AA. Methodology: TTHN, AA, EOA. Validation: AA, EOA, GP, and YZ. Result interpretation: TTHN, AA, EOA, DA, EAD, GP, YZ, and MB. Supervision: AA and EOA. Writing the original draft: TTHN, AA, and EOA. Critical revision and approval of the final version: TTHN, AA, EOA, DA, EAD, GP, YZ, MB, CIO, and VO.

## Data Availability

All data produced in the present work are contained in the manuscript and the supplementary section.

## Acknowledgements

We thank the authors of the original studies included in the systematic review

## Supplementary materials

Figure 1S. Funnel plot of incidence rates of Alzheimer’s disease among individuals with type 2 diabetes

Figure 2S. Funnel plot of adjusted relative risks of Alzheimer’s disease among individuals with type 2 diabetes

Table 1S – Search strategy

Table 2S – Quality rating of included studies

## List of abbreviations

AD: Alzheimer’s disease
aHR: Adjusted hazard ratio
aOR: Adjusted odds ratio
APOE: Apolipoprotein E
aRR: Adjusted relative risk
BMI: Body mass index
CI: Confidence interval
GRADE: Grading of Recommendations, Assessment, Development and Evaluations
HbA1c: Hemoglobin A1c
HR: Hazard ratio
ICD: International Classification of Diseases
IR: Incidence rate
MeSH: Medical Subject Headings
NHIRD: National Health Insurance Research Database
NINCDS-ADRDA: National Institute of Neurological and Communicative Disorders and Stroke–Alzheimer’s Disease and Related Disorders Association
NOS: Newcastle–Ottawa Scale
OR: Odds ratio
PI: Prediction interval
PRISMA: Preferred Reporting Items for Systematic Reviews and Meta-Analyses guidelines
PY: Person-years
REML: Restricted Maximum Likelihood
RR: Relative risk
T2D: Type 2 diabetes
UK: United Kingdom
US: United States

