## Supplementary figures and images for "Incidence and Risk of Alzheimer’s Disease in Individuals with Type 2 Diabetes: A Systematic Review and Meta-Analysis"

### Supplementary Figure 1

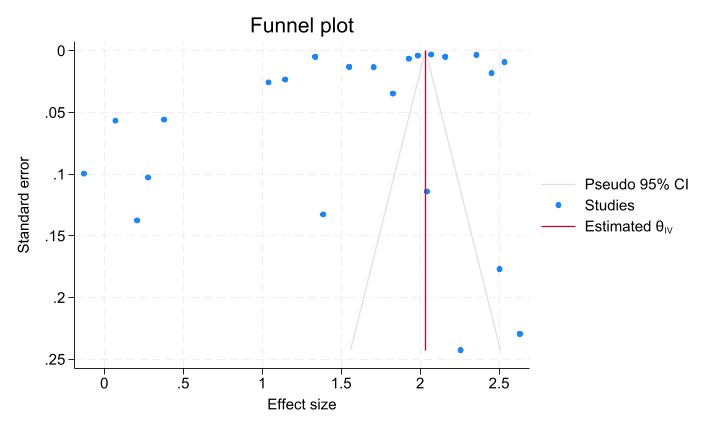

### Supplementary Figure 2

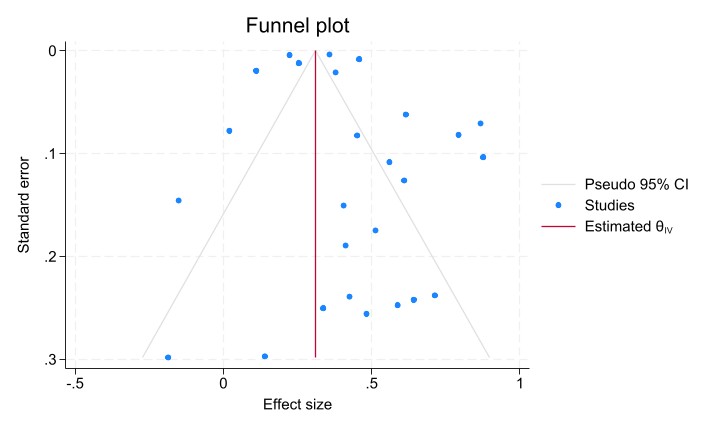
