## Supplementary Table 1 for "Incidence and Risk of Alzheimer’s Disease in Individuals with Type 2 Diabetes: A Systematic Review and Meta-Analysis"

**Table 1S – Search Strategy**

| # | Searches |
| --- | --- |
| 1 | Alzheimer’s OR Alzheimer’s disease OR Alzheimer Disease OR Alzheimer Syndrome |
| 2 | Diabetes Mellitus OR Diabetes OR Type 2 Diabetes Mellitus OR Adult-Onset Diabetes Mellitus OR Maturity-Onset Diabetes Mellitus OR Noninsulin-Dependent Diabetes Mellitus |
| 3 | Observational stud* OR Cohort stud* OR Case-control stud* OR Incidence Stud* OR Follow-up stud* OR Prospective stud* OR Retrospective stud* OR Cohort analysis OR Epidemiologic stud* OR Concurrent Stud* OR Cross-sectional study OR survey OR descriptive stud* |
| 4 | 1 and 2 and 3 |
| 5 | Limit 4 from inception to 30^th^ April 2026 |
