## Supplementary Table 2 for "Incidence and Risk of Alzheimer’s Disease in Individuals with Type 2 Diabetes: A Systematic Review and Meta-Analysis"

**Table 2S – Quality rating of included studies**

| Authors (Year) | Cohort study | Representativeness of the exposed cohort | Selection of the non-exposed cohort | Ascertainment of exposure | Demonstration that outcome of interest was not present at start of study | Comparability of cohorts on the basis of the design or analysis^(a)^ | Assessment of outcome | Was follow-up long enough for outcomes to occur | Adequacy of follow up of cohorts | Final score |
| --- | --- | --- | --- | --- | --- | --- | --- | --- | --- | --- |
|  | Cross-sectional study | Representativeness  of the study sample | Sample size | Assessment of the  Exposure^(a)^ | Assessment of  Outcome^(a)^ | Adjustment for  Confounder (s)^(a)^ | Assessment of confounder(s) |  |  |  |
|  | Case-control study | Adequate case definition | Representativeness of the cases | Selection of Controls | Definition of Controls | Comparability of cases and controls on the basis of the design or analysis ^(a)^ | Ascertainment of exposure | Same method of ascertainment for cases and controls | Non-Response rate |  |
| Leibson et al. 1997 | Cohort | * | * | * | * | * | * | * | * | 8/9 |
| Ott et al. 1999 | Cohort | * | * | * | * | * | * | * | * | 8/9 |
| Hassing et al. 2002 | Cohort | * | * | * |  | ** | * | * | * | 8/9 |
| Peila et al. 2002 | Cohort | * | * |  | * | ** | * | * | * | 8/9 |
| Borenstein et al. 2005 | Cohort | * | * |  | * |  | * | * | * | 6/9 |
| Chen et al. 2007 | Cross-sectional | * |  |  | ** | ** | * |  |  | 6/9 |
| Irie et al. 2008 | Cohort | * | * | * | * | ** | * | * | * | 9/9 |
| Xu et al. 2009 | Case-control | * | * | * | * | ** |  | * | * | 8/9 |
| Raffaitin et al. 2009 | Cohort | * | * | * | * | * | * | * | * | 8/9 |
| Al-Emam et al. 2010 | Cohort |  | * | * | * | * | * | * | * | 7/9 |
| Reitz et al. 2010 | Cohort | * | * |  | * | * | * | * | * | 7/9 |
| Cheng et al. 2011 | Cohort | * | * |  | * | ** |  | * | * | 7/9 |
| Huang et al. 2014 | Cohort | * | * | * | * | ** | * | * | * | 9/9 |
| Davis et al. 2017 | Cohort | * | * | * |  | * | * | * | * | 7/9 |
| Li et al. 2017 | Cohort | * | * | * | * | ** | * | * | * | 9/9 |
| Kuan et al. 2017 | Cohort | * | * | * | * | ** | * | * | * | 9/9 |
| Kim et al. 2017 | Cohort | * | * | * | * | ** | * | * | * | 9/9 |
| Nam et al. 2019 | Cohort | * | * | * | * | ** | * | * | * | 9/9 |
| Doney et al. 2019 | Cohort | * | * | * | * | * | * | * | * | 8/9 |
| Garfield et al. 2021 | Cohort | * | * |  | * | ** | * | * | * | 8/9 |
| Lee et al. 2021 | Cohort | * | * | * | * | * | * | * | * | 8/9 |
| Geng et al. 2022 | Cohort | * | * | * | * | ** | * | * | * | 9/9 |
| Han et al. 2022 | Cohort | * | * | * | * | ** | * | * | * | 9/9 |
| Kim et al. 2022 | Cohort | * | * |  | * | ** | * | * | * | 8/9 |
| Torrandell-Haro et al. 2022 | Cohort | * | * | * | * | ** | * | * | * | 9/9 |
| Park et al. 2022 | Cohort | * | * | * | * | ** | * | * | * | 9/9 |
| Lee et al. 2022 | Cohort | * | * | * | * | ** | * | * | * | 9/9 |
| Dove et al. 2023 | Cohort | * | * |  | * | ** | * | * | * | 8/9 |
| Zhou et al. 2023 | Cohort | * | * | * | * | ** | * | * | * | 9/9 |
| Wang et al. 2023 | Cohort | * | * |  | * | ** | * | * | * | 8/9 |
| Liu et al. 2024 | Cohort | * | * | * | * | ** | * | * | * | 9/9 |
| Seo et al. 2024 | Cohort | * | * | * | * | ** | * | * | * | 9/9 |
| Kim et al. 2024 | Cohort | * | * | * | * | ** | * | * | * | 9/9 |
| Merlini et al. 2025 | Cohort | * | * | * | * | ** | * | * | * | 9/9 |
| Zhang et al. 2025 | Cohort | * | * | * | * | ** | * | * | * | 9/9 |
| Jin et al. 2025 | Cohort | * | * | * | * | ** | * | * | * | 9/9 |
| Jun et al. 2026 | Cohort | * | * | * | * | ** | * | * | * | 9/9 |
| Khangura et al. 2026 | Cohort | * | * | * | * | ** | * | * | * | 9/9 |
| Dong et al. 2026 (a) | Cohort | * | * | * | * | ** | * | * | * | 9/9 |
| Dong et al. 2026 (b) | Cohort | * | * | * | * | ** | * | * | * | 9/9 |

(a): A maximum of 2 stars
